# Hidden Monogenic Diagnoses in Young-Onset Dialysis After Exclusion of Clinically Apparent Polycystic Kidney Disease

**DOI:** 10.64898/2026.10.17.26365780

**Authors:** Reiji Takami, Yoshinori Sato, Koichiro Higasa, Toshinori Imaizumi, Daisuke Tanaka, Daisuke Yabe, Nobuya Inagaki, Tsuyoshi Oshiro, Mayu Shimokawa, Keishu Kawanishi, Hidetoshi Ito, Fumihiko Koiwa

## Abstract

**Background:** Genomic reassessment may reveal clinically relevant diagnoses in patients with young-onset kidney failure despite established clinical diagnostic labels. We evaluated this approach after excluding clinically apparent polycystic kidney disease (PKD).

**Methods:** Among 1,139 patients receiving maintenance dialysis at participating Japanese facilities, 355 had initiated dialysis at age ≤50 years. After excluding 17 patients with clinically apparent PKD, 108 of 338 eligible patients underwent whole-genome sequencing through voluntary, nonrandom enrollment. Analysis included 465 nuclear candidate genes, mitochondrial DNA, and copy-number/structural variants. Clinically concordant diagnoses were adjudicated using integrated clinical and genetic evidence, with a separate pathogenic/likely pathogenic (P/LP)-supported subset.

**Results:** Diagnoses were identified in 10/108 participants (9.3%; 95% confidence interval, 5.1–16.2), comprising seven renal and three systemic/metabolic diagnoses. Nine diagnoses were P/LP-supported (8.3%; 95% confidence interval, 4.4–15.1). The remaining diagnosis was phenotype-based clinical Dent disease associated with a CLCN5 variant of uncertain significance. Diagnoses occurred across diabetic kidney disease, nephrosclerosis, chronic glomerulonephritis, unknown-cause, and other clinical categories. In a post hoc analysis excluding three participants with low target coverage, the integrated and P/LP-supported proportions were 9/105 (8.6%) and 8/105 (7.6%). Six exploratory findings were excluded from both diagnostic numerators. Gitelman syndrome was included as a clinically manifest renal disorder, although its contribution to kidney failure remained uncertain.

**Conclusions:** Integrated genomic assessment can uncover renal and systemic/metabolic diagnoses across conventional labels in young-onset kidney failure after exclusion of clinically apparent PKD. Nonrandom enrollment and variable sequencing coverage limit generalizability and interpretation of negative results.

## Introduction

Genomic sequencing can identify monogenic disorders underlying conventional diagnoses of chronic kidney disease and kidney failure.^1-3^ Although studies of young-onset kidney failure of undetermined etiology have demonstrated substantial diagnostic yields,^2^ the value of genomic reassessment also warrants evaluation in patients with established clinical diagnoses. Therefore, we applied whole-genome sequencing (WGS) to a Japanese young-onset dialysis cohort across preexisting diagnostic categories, excluding clinically apparent polycystic kidney disease (PKD), which is often recognized from imaging and clinical history.^4^

## Methods

Young-onset kidney failure was defined as kidney failure requiring dialysis initiation at age ≤50 years, consistent with a published genomic study.^2^ Among the 1,139 patients receiving maintenance dialysis at participating sites on June 1, 2024, 355 met this criterion; 338 eligible patients remained after exclusion of 17 with clinically apparent PKD (4.8%). Of these, 108 (32.0%) underwent WGS through voluntary, nonrandom enrollment between June 1, 2024, and June 10, 2025. The median dialysis-initiation age was 40.5 years (interquartile range, 33.8–45.0). Pre-WGS clinical categories were derived from the original medical records. Figure 1, Supplementary Figure S1, and Supplementary Table S5 show the cohort selection, clinical label distribution, and participant characteristics.

**Figure 1.**
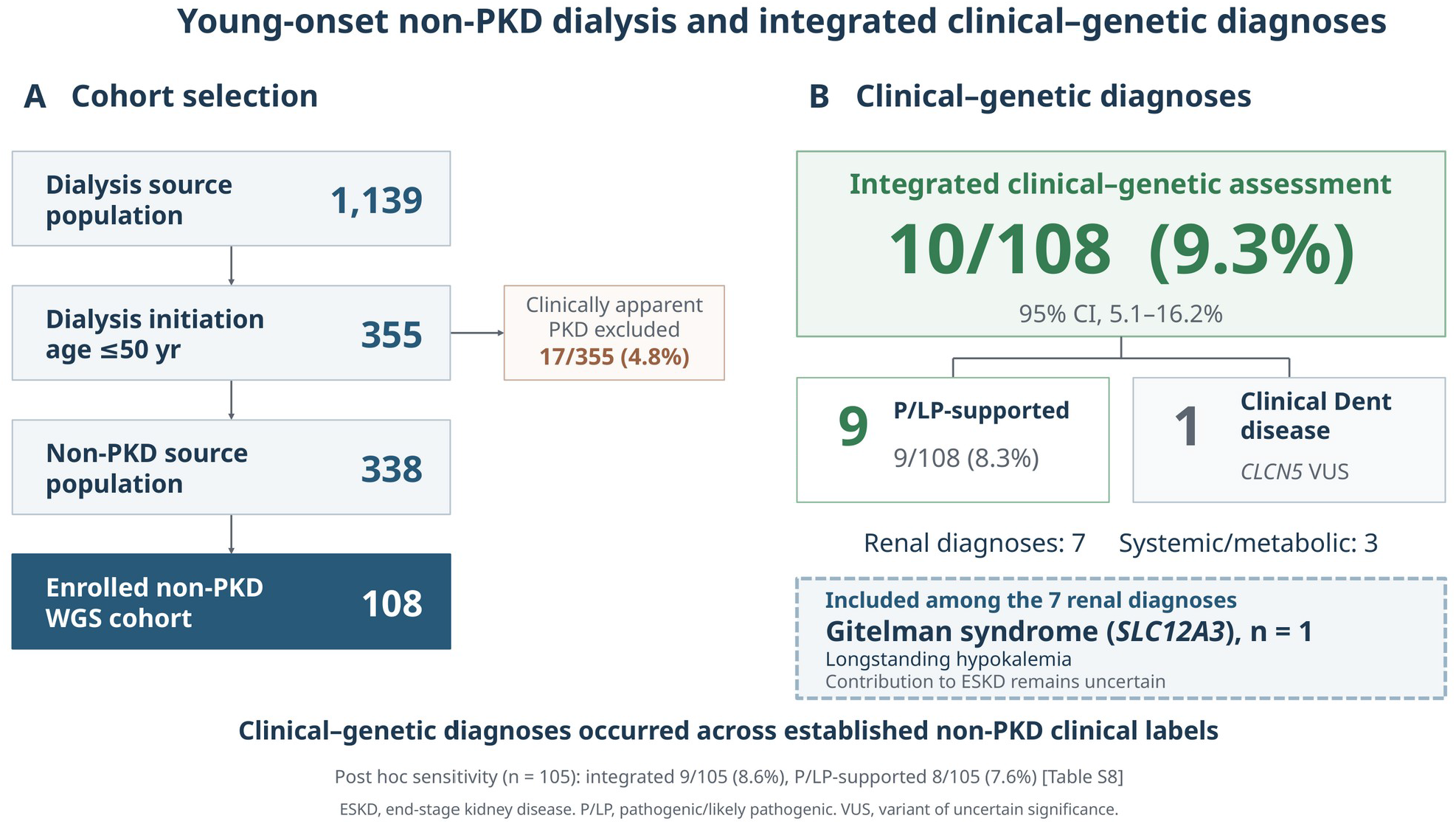
PKD-excluded young-onset dialysis cohort and integrated clinical–genetic diagnoses. The ten diagnoses comprise nine P/LP-supported diagnoses and one phenotype-based clinical Dent diagnosis with a *CLCN5* VUS; the latter is not molecular confirmation. Seven are renal diagnoses, including clinically manifest *SLC12A3*-related Gitelman syndrome, and three are systemic/metabolic diagnoses. The outcome captures clinically concordant disorders; it does not establish that each disorder was the sole cause of ESKD. The contribution of Gitelman syndrome to ESKD remained uncertain. The Wilson 95% confidence interval is shown for the primary outcome. The post hoc target-coverage sensitivity analysis excludes participants FJ-ESRD-002, FJ-ESRD-003, and FJ-ESRD-061, each with <50% of target bases covered at ≥10×. An additional analysis omitting the Gitelman diagnosis is provided in Supplementary Table S8. ESKD, end-stage kidney disease; P/LP, pathogenic/likely pathogenic; PKD, polycystic kidney disease; VUS, variant of uncertain significance; WGS, whole-genome sequencing.

Variant interpretation was conducted in accordance with the ACMG/AMP principles,^5^ relevant ClinGen recommendations, and genotype–phenotype concordance. The WGS analysis included 465 literature-derived nuclear candidate genes updated until July 2026, mitochondrial DNA, and copy-number/structural variants (CNVs/SVs) assessed using Manta, CNVkit, and exon-level read depth. The primary outcome comprised clinically concordant renal or systemic/metabolic diagnoses established through integrated clinical–genetic adjudication, without requiring attribution of kidney failure solely to the identified disorder. Pathogenic/likely pathogenic (P/LP)-supported diagnoses were distinguished from phenotype-based clinical diagnoses, and nondiagnostic findings were excluded. A post hoc sensitivity analysis excluded three participants with <50% of target bases covered at ≥10×. Analytical details, the gene list, and per-sample coverage are provided in the Supplementary Methods and Tables S7–S10.

## Results

Integrated clinical–genetic adjudication identified diagnoses in 10 of 108 participants (9.3%; 95% CI, 5.1– 16.2; Table 1 and Supplementary Table S4). Seven were renal diagnoses: two collagen IV-related disorders, *NPHP1* biallelic deletion, *NPHP3*-related disease, *LMX1B*-related disease, *SLC12A3*-related Gitelman syndrome, and clinical Dent disease. The Dent diagnosis was supported by childhood proteinuria predating diabetes, low-molecular-weight proteinuria, and renal calcification;^6^ *CLCN5* p.Leu693Arg remained a variant of uncertain significance (VUS), not molecular confirmation. The Gitelman syndrome diagnosis was supported by longstanding hypokalemia, although its contribution to kidney failure remains uncertain. The P/LP-supported subset comprised of 9/108 participants (8.3%; 95% CI, 4.4–15.1). In the 105-participant sensitivity analysis, the integrated and P/LP-supported proportions were 9/105 (8.6%) and 8/105 (7.6%), respectively (Supplementary Table S8).

**Table 1.**
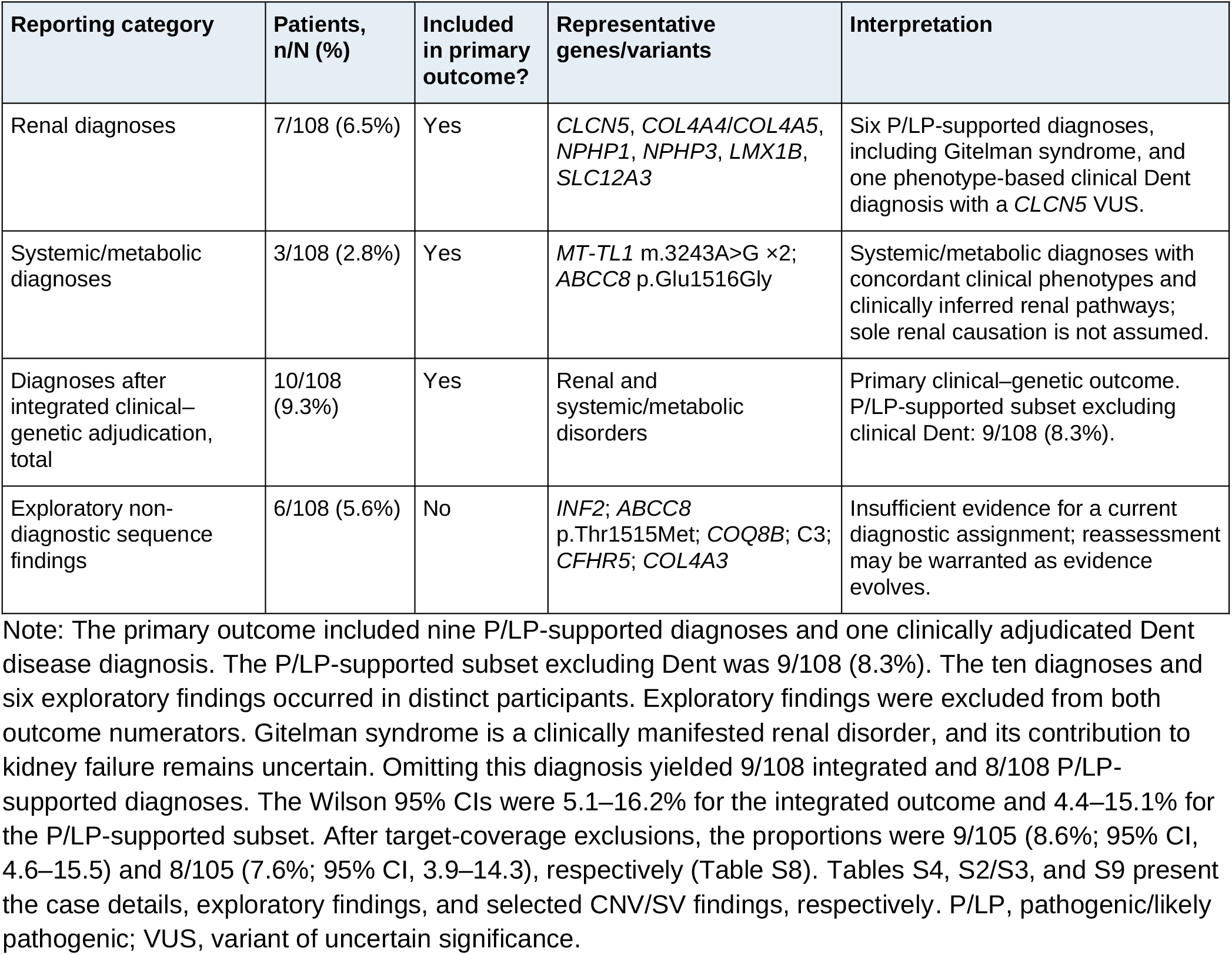
Summary of diagnoses and non-diagnostic findings after integrated clinical–genetic adjudication.

The three systemic/metabolic diagnoses comprised two MT-TL1 m.3243A>G-associated disorders and one ABCC8-related monogenic diabetes diagnosis based on the pathogenic classification of p.Glu1516Gly by experts. These findings informed the assessment of diabetes-mediated or other systemic contributions to kidney injury. Supplementary Table S4 provides individual clinical courses and adjudication evidence. Diagnoses occurred across the pre-WGS categories: diabetic kidney disease (4/33), nephrosclerosis (1/17), chronic glomerulonephritis (2/19), unknown cause (2/31), and other diagnoses (1/7), with no diagnoses in the vasculitis category (0/1). The P/LP-supported results by category are listed in Supplementary Table S6. No diagnoses associated with *PKD1, PKD2, UMOD*, or *HNF1B* were identified within the analytical scope.

The CNV/SV analysis supported the existing *NPHP1* biallelic deletion and established no additional kidney failure diagnoses among the 98 evaluable participants (Supplementary Tables S9 and S10). Six exploratory non-diagnostic sequence findings were excluded from the primary outcomes (Supplementary Tables S2 and S3). Among these, *INF2* p.Ser93Ala is of nephrologic interest, given the association of this gene with familial focal segmental glomerulosclerosis.^7^ However, variant-specific evidence was insufficient for diagnostic assignment. Therefore, a reassessment of these VUS findings may be required as variant-specific evidence and applicable phenotypic criteria evolve.

## Discussion

The contribution of this study is best understood alongside the differences in recruitment and diagnostic scope. Granhøj et al.^2^ reported WGS diagnoses in 26/114 patients (23%) with kidney failure of undetermined etiology by age 50. These diagnoses were reached after prior SNP-array and MUC1 testing, along with staged proband, family, and reanalysis approaches.^2^ Similarly, Liu et al.^3^ identified diagnoses by whole-exome sequencing in 23/317 patients (7.3%) from a broader dialysis cohort; PKD1 accounted for 11 of the 23 diagnoses. Our study extends this evidence to young-onset dialysis patients after exclusion of clinically apparent PKD, while retaining established non-PKD labels. Diagnoses across several clinical categories indicate the need for extended genomic evaluations beyond unexplained kidney failure. Recruitment and outcome definitions differ across studies. Therefore, diagnostic proportions are contextual rather than directly comparable.

Another strength of our approach is the joint assessment of renal, systemic, and metabolic disorders. A clinical diagnosis, such as that of diabetic kidney disease, may describe the mechanism of injury while leaving its upstream genetic cause unresolved. Similarly, identifying mitochondrial diseases can prompt the reassessment of renal involvement in diabetes and other organ manifestations. Integrating these findings with the clinical course can refine the etiological understanding without requiring replacement of the original renal diagnosis.

WGS provided a common platform for nuclear sequence, mitochondrial, and structural–variant analyses in this cohort. This breadth supports a genomic approach capable of evaluating renal and extrarenal etiologies beyond a narrowly defined pretest diagnosis. These findings illustrate the utility of WGS as an analytical foundation for phenotype-informed reassessment.

The small, non-randomly enrolled prevalent dialysis cohort limits precision and generalizability. Variable WGS coverage and mapping constraints limited the exclusion of genetic diseases from negative results, particularly in the three low-coverage participants who were also excluded from the CNV interpretation. Supplementary Tables S8–S10 show the per-sample coverage, analysis-specific exclusions, and sensitivity results.

Genomic evaluation should be considered for young-onset kidney failure, irrespective of the existing clinical diagnostic labels. Even after clinically apparent PKD is excluded, an integrated WGS-based assessment can uncover the renal and systemic or metabolic diagnoses underlying conventional labels. Therefore, an established diagnosis of diabetic kidney disease, nephrosclerosis, or glomerulonephritis should not preclude a genetic evaluation.

## Supporting information

Supplementary methods, figures, tables, references, and sequencing quality metrics.

## Data Availability

All data produced in the present study are available upon reasonable request to the authors

## Ethics Statement

This study was approved by the Showa Medical University Ethics Committee (approval no. 2024-070-A). Initial ethics approval and permission to commence the study were obtained on May 25, 2024, before informed consent and sample collection began on June 1, 2024. Ethical approval was obtained from Kyoto University on September 28, 2026 (approval no. G1029-1). Written informed consent for genomic analysis was obtained from all the 108 enrolled participants.

## Author Contributions

RT, YS, HI, and FK designed the study. RT, YS, TO, and KK obtained informed consent. RT, YS, and TO collected the DNA samples. TO, MS, and KK collected clinical data. MS and KK examined pathological specimens. KH performed the bioinformatic analyses. YS, TI, DT, DY, and NI performed the genetic interpretation. HI and FK coordinated the ethical approval for the study. YS, KH, TI, DT, DY, NI, HI, and FK contributed to manuscript preparation.

## Data Availability Statement

Individual-level genomic data cannot be publicly shared due to privacy and consent restrictions. Deidentified aggregate data supporting the findings are available from the corresponding author upon reasonable request and after appropriate approval.

## Acknowledgments

We would like to thank the patients who participated in this study. We also thank Drs. Chiaki Kumata, Kae Ito, and Kanji Shishido of the Kawasaki Clinic, as well as the staff of the dialysis facilities of the Zenjinkai Group, Miho-no-Mori Clinic (Akaedakai), and Koshikawa Memorial Yokohama Kidney Clinic (Sowakai) for their cooperation in clinical data collection.

## Supplementary Material

Supplementary Material: Supplementary Methods, Supplementary Figure S1, Supplementary Tables S1– S10, and Supplementary References.

## Declaration of generative AI and AI-assisted technologies in the manuscript preparation process

During the preparation of this work, ChatGPT (OpenAI) was used for English-language editing, reference organization, table formatting, and preparation of reproducible figure-building codes. Claude (Anthropic) was used to develop analysis scripts for coverage and CNV/SV analyses and summarize their results. Scientific interpretation and final variant classification were the responsibility of the study investigators.

## References

1. Groopman EE, Marasa M, Cameron-Christie S, et al. Diagnostic utility of exome sequencing for kidney disease. N Engl J Med. 2019;380:142–151. doi:10.1056/NEJMoa1806891.

2. Granhøj J, Aagaard MM, Pedersen KV, et al. Diagnostic yield of whole-genome sequencing in patients with kidney failure of undetermined etiology at age 50 years or younger. Kidney Int Rep. 2025;10(11):3984–3997. doi:10.1016/j.ekir.2025.09.001.

3. Liu ZY, Zhang YL, Li Y, et al. Screening for genetic kidney diseases in a dialysis cohort via exome sequencing. Clin Kidney J. 2026;19(2):sfag015. doi:10.1093/ckj/sfag015.

4. Pei Y, Obaji J, Dupuis A, et al. Unified criteria for ultrasonographic diagnosis of ADPKD. J Am Soc Nephrol. 2009;20:205–212.

5. Richards S, Aziz N, Bale S, et al. Standards and guidelines for the interpretation of sequence variants. Genet Med. 2015;17:405–424.

6. Bökenkamp A, Ariceta G, Böckenhauer D, et al. Dent disease: clinical practice recommendations. Nephrol Dial Transplant. 2025;40:852–864.

7. Boyer O, Benoit G, Gribouval O, et al. Mutations in INF2 are a major cause of autosomal dominant focal segmental glomerulosclerosis. J Am Soc Nephrol. 2011;22:239–245.

