## Supplementary methods, figures, tables, references, and sequencing quality metrics. for "Hidden Monogenic Diagnoses in Young-Onset Dialysis After Exclusion of Clinically Apparent Polycystic Kidney Disease"

**Study population.** Young-onset kidney failure was operationally defined as a diagnosis requiring dialysis initiation at ≤50 years of age, consistent with the age threshold used in a published genomic study of kidney failure of undetermined etiology.[S1] This refers to age at dialysis initiation, not age at enrollment. We enumerated the patients receiving maintenance dialysis at the participating facilities on June 1, 2024. Patients with clinically apparent polycystic kidney disease (PKD), defined by characteristic bilateral polycystic kidney morphology and a compatible clinical history, were excluded. Among the 1,139 patients, 355 had initiated dialysis at ≤50 years of age; 338 non-PKD patients remained following exclusion of 17 with clinically apparent PKD. Of these, 108 (32.0%) underwent whole-genome sequencing (WGS) and integrated clinical–genetic adjudication. Recruitment by nephrologists was voluntary and non-random. Written informed consent and samples were obtained between June 1, 2024, and June 10, 2025.

**Participant characteristics and descriptive analysis.** The participant characteristics are summarized in Supplementary Table S5. Continuous variables are reported as medians and interquartile ranges (25th–75th percentiles; inclusive linear interpolation), whereas categorical variables are expressed as counts and percentages. Ages at dialysis initiation and on the June 1, 2024 census date were available for all 108 participants. Participant sex was confirmed by the study investigators and was concordant with the sex chromosome read-depth inference (Supplementary Table S10). Study and technical sample identifiers were reconciled using an investigator-confirmed crosswalk. In addition, withdrawal and clinically apparent PKD samples were excluded from the final participant summaries. Cohort-level family histories were not summarized because ascertainment and documentation were considered insufficiently reliable for aggregate analysis; individually verified histories were retained in case descriptions. The dialysis duration was not estimated based on integer age differences. Furthermore, detailed comparisons with nonparticipants were not performed because some facilities did not authorize the provision or publication of these data within the approved study arrangements.

**Diagnostic proportions and sensitivity analysis.** The diagnostic proportions were reported with Wilson 95% confidence intervals. In total, 108 participants were included in the primary analysis. The post hoc sensitivity analysis excluded participants in whom <50% of the target bases across the coding and splice-flanking regions of the 465 nuclear genes were covered at ≥10×. Three participants (FJ-ESRD-002, FJ-ESRD-003, and FJ-ESRD-061) met this criterion, resulting in 105 participants. This criterion was applied irrespective of diagnostic status. The IGV-reviewed *COL4A4* detection in FJ-ESRD-061 remained in the primary analysis but was excluded from the sensitivity analysis together with the participant. The integrated clinical–genetic diagnoses and the P/LP-supported subset excluding clinical Dent disease are shown separately in Supplementary Table S8. This analysis assessed changes in observed proportions, not false-negative rates or equivalence with higher-depth WGS.

**Pre-WGS clinical categories.** The enrolled participants were grouped according to the diagnoses documented in the original clinical records. This grouping was conducted separately from subsequent clinical–genetic etiologic reassessment. Participant FJ-ESRD-048 was assigned to chronic glomerulonephritis (CGN) because IgA nephropathy was the principal historical diagnosis in two biopsy reports, although a later review raised uncertainty regarding the dominant renal mechanism. IgA nephropathy, FSGS, and lupus nephritis were grouped as CGN, whereas interstitial nephritis and CAKUT were grouped as Others. DMN and BNS corresponded to diabetic kidney disease and nephrosclerosis, respectively; the RPGN-labelled participants retained the vasculitis category. The enrolled counts were 33, 17, 19, 31, 1, and 7 participants for diabetic kidney disease, nephrosclerosis, CGN, unknown cause, vasculitis, and other conditions, respectively. Supplementary Table S6 shows category-specific diagnostic proportions.

**Ethics and consent.** This study was approved by the Showa Medical University Ethics Committee (approval no. 2024-070-A). Initial ethics approval and permission to commence the study were obtained on May 25, 2024, before informed consent and sample collection began on June 1, 2024. Ethical approval was obtained from Kyoto University on September 28, 2026 (approval no. G1029-1). Written informed consent for genomic analysis was obtained from all 108 enrolled participants.

**Whole-genome sequencing and bioinformatic analysis.** Genomic DNA was extracted from peripheral blood leukocytes. PCR-free libraries were prepared using the TruSeq DNA PCR-Free Library Preparation Kit and sequenced on a NovaSeq X Plus platform with 150-bp paired-end reads. The reads were aligned to hg19 (GRCh37) using BWA v0.7.13-r1126. Single-nucleotide variants and small insertions/deletions were called using GATK v3.8-1-0-gf15c1c3ef with the GATK default filtering criteria. The MUC1 coding variable-number tandem-repeat (VNTR) region was screened using VNtyper;[S2] no positive MUC1 VNTR findings were identified among the 108 participants. Additional coverage, CNV/SV, and mitochondrial analyses are described below.

**Target-region coverage.** The coverage for each of the 465 nuclear candidate genes was assessed using mosdepth v0.3.7[S3] over the coding exons of the longest coding transcript in the UCSC hg19 refFlat annotation. No mapping quality filter was applied. In addition, duplicate, secondary, and QC-failed reads were excluded by default. Coding intervals were extended by 20 bp on each side, and overlapping intervals within each gene were merged to yield 1,490,604 target bases. Per-sample mean depth and fractions covered at ≥1×, ≥10×, ≥20×, and ≥30× were calculated. Autosomal and X-linked targets were evaluated separately, with sex-specific summaries for X-linked genes. Among the final 108 participants, the median of sample-level genome-wide mean depths was 13.7× (range, 3.8–61.1×), and the median target-region mean depth was 14.2× (5.7–59.5×). The median fractions covered at ≥10× and ≥20× across all targets were 86.9% and 10.5%, respectively; the median autosomal ≥10× fraction was 89.0%. The ≥10× target fractions were 40.94%, 16.86%, and 40.08% for FJ-ESRD-002, FJ-ESRD-003, and FJ-ESRD-061, respectively; the next-lowest value was 78.67%. These three participants constituted the post hoc sensitivity exclusion set. Supplementary Table S10 presents the participant-level QC values.

**Copy-number analysis.** Read-depth copy-number analysis was performed using CNVkit v0.9.10[S4] in whole-genome mode using 10-kb bins, a pooled sex-adjusted reference, circular binary segmentation, and sex-aware threshold calling. The pooled CNVkit reference was built from the genomes sequenced for this project, excluding the two lowest-depth samples (FJ-ESRD-002 and FJ-ESRD-003). CNVkit segments ≥20 kb spanning ≥3 bins were reviewed. A batch-matched reference was constructed from 83 other genomes for samples prepared in a separate library batch. Exon-level mean depths were normalized to each sample's autosomal median and then to the corresponding cohort median; same-sex comparisons were used for sex chromosome loci. Relative exon depths are expressed as log2 ratios.

**Structural-variant analysis.** Structural variants were called using Manta v1.6.0[S5] in the diploid mode, and chromosome-level outputs were combined. PASS calls with overlapping candidate gene spans were annotated with carrier counts within the analyzed cohort and in 81 unrelated in-house comparison genomes. Manta PASS calls with a within-cohort carrier frequency ≤5% were reviewed. Calls were grouped for carrier counting using a 50% reciprocal overlap. Candidate deletions and duplications were reviewed for concordant CNVkit segments and exon-level read depth changes. Exon-level abnormalities were not interpreted as diagnostic CNVs in isolation.

**Analysis-specific quality assessment.** Samples were excluded from the CNV/SV interpretation when the genome-wide mean depth was <10× or the median absolute deviation of the CNVkit bin-level log2 ratios exceeded 0.15. In the final 108-participant cohort, 98 participants were evaluable—92 using the standard reference and six using a batch-matched reference. Ten participants were excluded from the CNV/SV interpretation rather than treated as negative: three with low coverage (FJ-ESRD-002, FJ-ESRD-003, and FJ-ESRD-061) and seven with excessive read-depth noise (FJ-ESRD-048, FJ-ESRD-050, FJ-ESRD-051, FJ-ESRD-052, FJ-ESRD-053, FJ-ESRD-062, and FJ-ESRD-065). Supplementary Table S10 shows the final per-sample QC classification. This assessment was separate from both SNV/indel and mitochondrial interpretations and the 105-participant target-coverage sensitivity subset. The *COL4A4* p.Gly1448Ala SNV in FJ-ESRD-061 remained in the primary analysis following the manual IGV review.

**Reassessment of the *NPHP1* deletion.** The previously identified biallelic NPHP1 deletion was reassessed using CNVkit segments and read-depth profiles across all 20 coding exons. The agreement between these analyses was considered concordant computational support from the same WGS data rather than orthogonal experimental validation. In FJ-ESRD-029, CNVkit identified a copy-number-0 segment at hg19 chr2:110,851,326–110,981,323 (log2 ratio −7.04), and all 20 coding exons had zero depth. Manta did not call this deletion. The CNVkit segment boundaries did not establish exact deletion breakpoints.

**Mitochondrial analysis.** For mitochondrial analysis, reads were realigned to the T2T-CHM13 v2.0 reference, which contains the revised Cambridge Reference Sequence. They were subsequently processed using MitoHPC (version 20240207).[S6] Processing included the removal of NUMT-derived reads, Mutect2-based heteroplasmy calling with a 3% threshold, and haplocheck contamination assessment.[S7] Mean mtDNA depth was obtained from the MitoHPC output and additionally assessed with mosdepth on chrM BAM files. The median of per-sample MitoHPC mean coverages across the 108 participants was 1,005.79× (range, 422.29–1,883.07×). Mean mtDNA coverage was 1,763.28× in FJ-ESRD-048 and 714.21× in FJ-ESRD-056; these sample-level means are distinct from variant-site read depths. m.3243A>G was detected at 17.5% in FJ-ESRD-048 and 19.0% in FJ-ESRD-056, and the same findings were retained at 5% and 10% heteroplasmy thresholds. mtDNA coverage in the three participants with low nuclear target coverage was 497.24–1,613.93×; the nuclear coverage finding was not treated as invalidating their mitochondrial analysis.

**Nuclear candidate-gene list.** The nuclear candidate-gene list was curated based on published renal gene lists and previous genetic-testing studies.[S8–S11] Genes were excluded based on congenital-onset disease during curation, and additional kidney disease–related genes were incorporated from relevant reports until July 2026. The reporting list comprised 465 unique nuclear genes (Supplementary Table S7). July 2026 denotes the reported literature update endpoint, and not a prospectively fixed panel at recruitment. The HGNC identifiers and hg19 gene span annotations were supplied separately from the selection provenance. These spans include introns and untranslated regions (UTRs); they are not CDS/splice-site coverage masks. Mitochondrial analysis and MUC1 coding-VNTR screening are described separately.

**Read-level confirmation.** Using the existing procedure of the bioinformatics team, potentially reportable variants with quality scores below 100 were flagged for additional read-level reviews. *COL4A4* p.Gly1448Ala and *CLCN5* p.Leu693Arg calls were manually reviewed in IGV. This review used the original WGS alignments and was not an orthogonal validation study. The threshold was a review trigger, not an automatic exclusion criterion; the respective LP and variant of uncertain significance (VUS) classifications remained unchanged.

**Selected negative findings.** No SNV or small indels meeting the diagnostic criteria were identified in *PKD1*, *PKD2*, *UMOD*, or *HNF1B* among the 108 non-PKD participants. Additionally, a manual IGV/read-depth review of *PKD1* and *HNF1B* did not reveal any diagnostic findings. These negative findings reflect the scope of the analyses performed; they do not establish the comprehensive exclusion of all sequences or structural variants of these genes. No additional diagnosis explaining kidney failure was established from the CNV/SV analysis of the 98 evaluable participants (Supplementary Table S10). Selected findings are summarized in Supplementary Table S9. A review of the available coding-variant table did not identify a second sequence variant in *BBS5* in duplication carriers FJ-ESRD-046/FJ-ESRD-054 or in AGT in deletion carrier FJ-ESRD-107. This review does not establish complete gene-level callability. FJ-ESRD-067 had an approximately 40-kb intronic *PKHD1* gain that did not overlap the coding exons and had no supporting Manta call. However, this signal did not establish an additional diagnosis (Supplementary Table S9).

**Variant interpretation and targeted review.** Variants were interpreted using the ACMG/AMP criteria,[S12] relevant ClinGen-related recommendations, population-frequency databases, in silico predictions, inheritance, published literature, and genotype–phenotype concordance. The final classification of *COL4A4* NM_000092:c.4343G>C (p.Gly1448Ala) was confirmed as likely pathogenic using PM1, PP3_Moderate, PM2_Supporting, and PP4. The interpretation incorporated Alport-specific guidance[S13] and calibrated the computational evidence for REVEL (score, 0.861).[S14] Renal histopathology and family history of hematuria and kidney impairment were concordant. Family segregation analysis was not performed, and de novo status was not established; PP1, PS2, and PM6 were not used for this variant. The location of COQ8B chr19:41220956 A>T (hg19) within the 5′ UTR of NM_001142555 was manually reviewed in IGV. RNA-based functional analysis was not performed, and its functional consequences remain unverified (Supplementary Table S3).

**Expert adjudication.** The selected variants were adjudicated by the study team at the patient level and, when appropriate, by experts in kidney pathology, inherited kidney disease, mitochondrial disease, or monogenic diabetes. Coded variant information and minimal clinical summaries were reviewed by diabetes experts for monogenic diabetes-related variants. Direct identifiers were not shared for expert adjudication. ABCC8 classifications and ACMG/AMP evidence strengths were supplied by the Monogenic Diabetes Expert Committee and were reproduced as committee adjudications; they were distinguished from public database assertions (Supplementary Table S2).

**Clinical chronology and renal pathways.** Supplementary Table S4 presents the expanded case-level information. In FJ-ESRD-044, diabetes onset at age 14 was distinguished from suspected MODY recorded in the referral letter at age 15. Autoantibody negativity, initial sulfonylurea response, insulin initiation, a three-generation diabetes history, retinopathy, and the renal course are described. Neither the patient nor family members reported a history of neonatal hypoglycemia. The referral letter described initially preserved insulin secretion with suspected later depletion, and persistently elevated HbA1c levels before kidney failure. Table S4 shows the updated glycemic and medication histories, together with unavailable assay details. For FJ-ESRD-048, neurological and bladder/bowel dysfunction, biopsy findings of IgA nephropathy and reflux-related injury, medication-free diabetes control, and confirmed maternal m.3243A>G variant supported the reassessment of the systemic phenotype. Severe cardiomyopathy, insulin-dependent diabetes, retinopathy, hearing loss, and short stature were described for FJ-ESRD-056; the mother declined genetic testing, and the proband had not undergone kidney biopsy. The proposed renal pathways are clinical inferences and do not prove that the variants are the sole causes of kidney failure.

**Reporting categories.** The primary outcome was renal or systemic/metabolic disorders established through integrated clinical–genetic adjudication concordant with the observed clinical phenotype. Variant classification, patient-level diagnosis, and contribution to kidney failure were assessed separately, and sole causation of kidney failure was not required. P/LP-supported diagnoses were distinguished from the phenotype-based clinical Dent diagnosis associated with a *CLCN5* VUS, which was not molecularly confirmed. The P/LP-supported subset excludes the Dent case. During the final clinical review, the detection of homozygous pathogenic *SLC12A3* in FJ-ESRD-059 was included as a clinical manifestation of Gitelman syndrome after long-standing hypokalemia and additional clinical findings were documented. However, its role in kidney failure remains unclear. An additional post hoc analysis omitting this diagnosis from the numerators with unchanged denominators is presented in

Table S8. Six sequence findings were retained for transparent exploratory reporting, with supporting observations and reasons for the nondiagnostic status stated individually in Table S3. No graded candidate categories or rankings of VUS toward likely pathogenicity were used. In particular, *ABCC8* p.Thr1515Met remained a committee-classified VUS. Earlier conditional correspondence did not constitute committee confirmation of a candidate diagnosis. All six findings were excluded from the primary outcome, occurred in distinct participants, and did not overlap with the ten diagnoses shown in Table S4. The selected non-diagnostic CNV/SV findings are reported separately in Table S9 and were not included in the sequence-finding counts.

Reassessment of unresolved findings. The current nondiagnostic status does not preclude later reinterpretation. For *INF2* p.Ser93Ala, additional well-characterized clinical reports, segregation studies, and functional evidence may address the lack of variant-specific support. For *ABCC8* p.Thr1515Met, the committee noted that the LP could be reached if PP4 was appropriately assigned in addition to PM1, PM2, and PP3. This conditional framework is distinguished from the current assessment, which does not assign PP4. In addition, neither future reclassification nor the probability of upgrading is established.

**Result return.** P/LP or otherwise clinically useful findings may be considered for return after genetic counseling according to participant preference and institutional policy. The Monogenic Diabetes Committee considered the FJ-ESRD-044 findings suitable for disclosure; the present report does not establish that disclosure has occurred. Exploratory non-diagnostic findings are not returned as confirmed diagnoses unless subsequent evidence establishes clinical validity and appropriate ethical procedures are completed.

**Analytical scope.** This study was not designed to compare WGS with whole-exome sequencing. Thus, genome-wide deep intronic and regulatory variants were not systematically incorporated into diagnostic adjudications. Variable sequencing depths, analysis-specific CNV/SV exclusions, and repetitive region mapping limitations restrict negative findings. In particular, small exon-level changes and changes in highly homologous regions cannot be fully assessed using these short-read methods. The concordant CNVkit and exon-level read-depth results used the same sequencing data and are not orthogonal laboratory validations. In the three low-target-coverage participants, negative nuclear sequence findings did not confidently exclude variants in known disease genes. Individually reviewed positive findings were retained in the primary analysis. The three participants were excluded from the CNV/SV interpretation and target-coverage sensitivity analysis.

**Figure preparation.** Supplementary Figure S1 was prepared from supplied source-population aggregates and the reconciled pre-WGS categories of enrolled participants using editable native PowerPoint charts. Percentages use the corresponding cohort denominator. Source-population counts were retained as originally supplied and were not re-adjudicated. Aggregate data and the revision code are retained in the study records. AI-assisted code preparation is disclosed in the main manuscript.

**Supplementary Figure S1. Selection and clinical diagnostic composition of the non-PKD WGS cohort.**

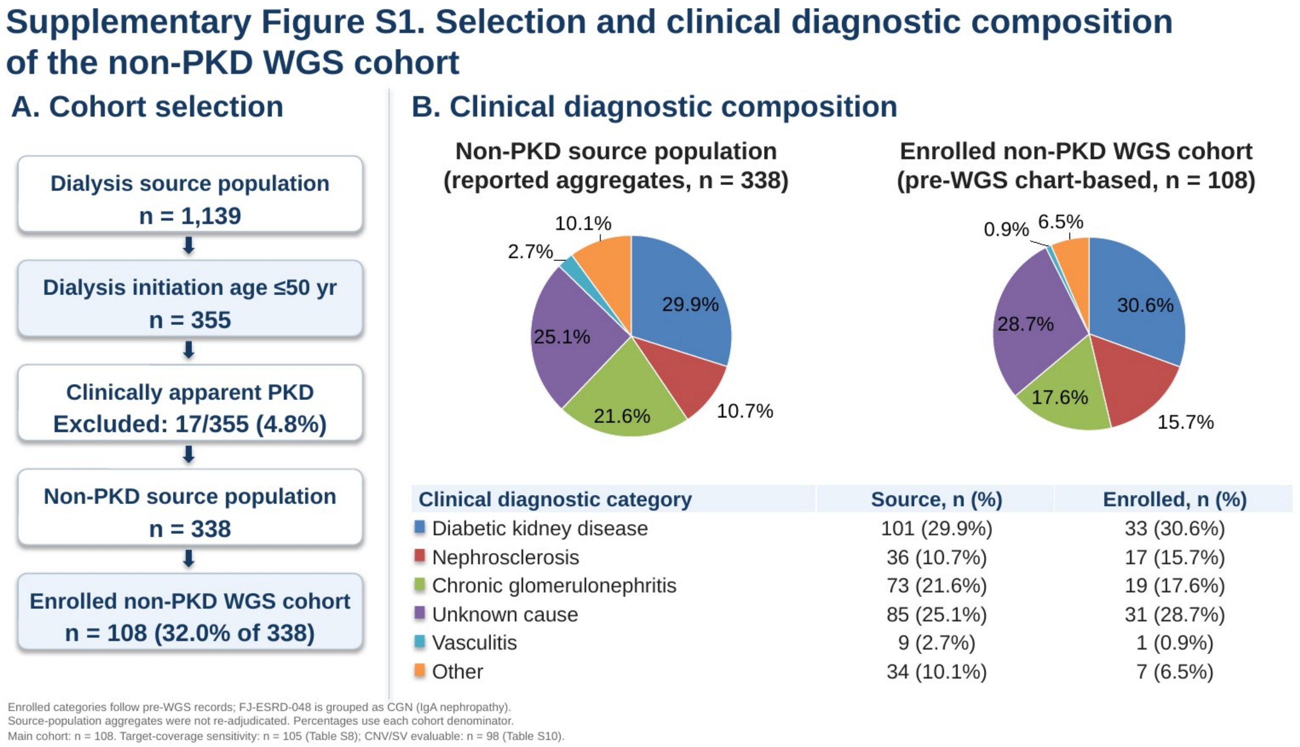


Panel A shows the cohort-selection process. Panel B shows the originally supplied source population counts and reconciled pre-WGS categories of the enrolled cohort. FJ-ESRD-048 was grouped as CGN (historical IgA nephropathy); subsequent etiologic uncertainty did not change the pre-WGS category. The source-population aggregates were not readjudicated. The census date was June 1, 2024, whereas consent and samples were obtained from June 1, 2024, to June 10, 2025. Percentages were calculated using each cohort denominator. The full clinical cohort (n = 108) is shown, and the separate post-hoc target-coverage sensitivity analysis included 105 participants (Supplementary Table S8). The CNV/SV-evaluable subset comprised 98 participants (Supplementary Table S10). CGN, chronic glomerulonephritis; PKD, polycystic kidney disease; WGS, whole-genome sequencing.

### Supplementary Table S1. Operational classification framework for retained findings

| Category | Basis of adjudication | Included in primary outcome? | Comment |
|---|---|---|---|
| Renal diagnosis | P/LP-supported renal disease, or a separately identified phenotype-based clinical diagnosis | Yes | Six P/LP-supported diagnoses, including clinically manifest Gitelman syndrome, and one clinical Dent diagnosis. The *CLCN5* VUS is not molecular confirmation. Gitelman's contribution to kidney failure remains unclear. |
| Systemic/metabolic diagnosis | Systemic or metabolic disease supported by P/LP findings and concordant phenotype, with a clinically plausible link to kidney injury | Yes | Includes MT-TL1-related disease and expert-adjudicated *ABCC8*-related diabetes. Syndrome diagnosis and renal causal pathway are assessed separately. |
| Exploratory non-diagnostic finding | Reported gene/phenotype, inheritance, or variant-level observations, with insufficient evidence for a patient-level diagnosis | No | *ABCC8* p.Thr1515Met and the five other sequence findings in Table S3. Retention does not imply greater likelihood of pathogenicity than other VUS. |

The primary outcome represents diagnoses after integrated clinical–genetic adjudication rather than molecular-only diagnostic yield. Clinically adjudicated Dent diagnosis is distinguished from P/LP-supported diagnoses and considered in the context of Dent disease recommendations.[S15] *COL4A4* p.Gly1448Ala is classified as likely pathogenic, not as a VUS exception.

### Supplementary Table S2. *ABCC8* adjudication summary

| Variant | Current committee classification | Study reporting category | Reason |
|---|---|---|---|
| FJ-ESRD-044 p.Glu1516Gly | Pathogenic | Systemic/metabolic diagnosis | Committee: PS3_moderate, PS4, PM1, PM2, PM5. Published inactivation/HI evidence, *NBD2* location, absence from gnomAD/ToMMo, and a pathogenic same-residue substitution support the committee classification. Non-obese diabetes, a three-generation family history, and initial sulfonylurea response support patient-level concordance. ClinVar 1338496: LP. See Supplementary Methods, S2 details, and S4. |
| FJ-ESRD-074 p.Thr1515Met | VUS | Exploratory non-diagnostic finding | Committee: PM1 + PM2 + PP3 (REVEL 0.886); PP4 not applied. Corrected PM2: gnomAD v4 joint grpmax FAF95 $1.83 \times 10^{-6}$ (threshold $\leq 3 \times 10^{-6}$). ClinVar 1506182: P/LP, non-MD-VCEP submitters. LP could be reached if PP4 were justified under an applicable framework; this is conditional. Current classification remains VUS, without a committee-confirmed candidate diagnosis. |
| p.Arg298Cys | VUS with BS1_strong | Not retained after *ABCC8* review | Population frequency provides strong benign evidence. |
| p.Arg1182Pro | VUS | Not retained after *ABCC8* review | No functional or segregation data; limited *ABCC8*-MODY-specific phenotype support. |

Supplementary Table S2 shows the Monogenic-Diabetes Expert Committee's *ABCC8* classifications and the supplied evidence; it is not an inventory of all WGS VUS. ClinVar assertions and ClinGen Monogenic Diabetes Variant Curation Expert Panel (MD-VCEP) reviews were distinguished from committee's adjudication. The PM2 correction for p.Thr1515Met does not change its VUS classification or its exclusion from the primary outcome.

***ABCC8* p.Glu1516Gly (FJ-ESRD-044).** The committee classified NM_000352.6:c.4547A>G as pathogenic using PS3 (moderate; published inactivation assays), PS4 (strong; a previously reported neonatal HI case), PM1 (moderate; NBD2), PM2 (moderate; absent from gnomAD and ToMMo in the committee review), and PM5 (moderate; another pathogenic substitution at the same residue). Boodhansingh et al.[S16] reported impaired channel activation and HI-associated observations; MacMullen et al.[S17] reported the same-residue p.Glu1517Lys substitution in their numbering. These strengths and the final classification are reproduced from the committee findings, rather than independently recalculated. The cited functional and clinical observations are primarily related to HI. Patient-level monogenic diabetes adjudication also considered non-obesity, diabetes across three generations, and initial sulfonylurea responsiveness. A direct renal effect was not inferred.

**Transcript correspondence.** Boodhansingh et al.[S16] used GenBank L78224, which includes an alternatively spliced sequence near exon 17 and encodes one additional amino acid. Their c.4550A>G, p.Glu1517Gly corresponds to NM_000352.6:c.4547A>G, p.Glu1516Gly. ClinVar Variation ID 1338496 lists this allele as likely pathogenic, which is distinct from the committee's pathogenic classification in this study.

***ABCC8* p.Thr1515Met (FJ-ESRD-074).** The committee retained NM_000352.6:c.4544C>T as the VUS using PM1 (moderate; NBD2), PM2 (moderate), and PP3 (supporting; REVEL 0.886). The corrected PM2 assessment used gnomAD v4 joint grpmax FAF95 of $1.83 \times 10^{-6}$, which is below the committee-cited threshold of $3 \times 10^{-6}$; the non-Finnish European AF was $8.813 \times 10^{-6}$, with absence reported in Japanese/East Asian data. These frequency values and criterion assignments are transcribed from the committee clarification. The prior June 28 assessment did not apply PM2, whereas the updated assessment applied PM2 but retained the VUS. PP4 was not applied, because the committee no longer regarded the general MODY referral criteria alone as sufficient for genes without a specified MD-VCEP phenotype. ClinVar Variation ID 1506182 contains P/LP assertions from non-MD-VCEP submitters; the committee does not adopt these assertions as expert panel classifications. An earlier conditional discussion on LP under a framework that assigned PP4 was not a committee-confirmed candidate diagnosis.

**Functional and phenotypic context.** Diabetes can occur with both activating and inactivating *ABCC8* variants through different mechanisms.[S18] In the Japanese JDS cohort, three of four *ABCC8*-MODY cases had a personal or family history of neonatal hypoglycemia.[S19] Clemente et al.[S20] documented later diabetes in a cohort with confirmed preceding CHI. Similarly, Kapoor et al.[S21] reported diabetes/gestational diabetes in carriers without a reported hypoglycemia history. Such histories have variable ascertainment and do not exclude unrecognized earlier HI. Although Scala et al.[S18] provide functional support for loss-of-function variants in MODY, individual prior-HI histories in their analyzed cohort were not established as negative. No pooled count of "MODY without CHI" is inferred from these heterogeneous reports. These observations provide a disease-level context and do not upgrade p.Thr1515Met or establish a prior CHI course in patients with FJ-ESRD-044.

**Database sources.** ClinVar: https://www.ncbi.nlm.nih.gov/clinvar/variation/1338496/ and https://www.ncbi.nlm.nih.gov/clinvar/variation/1506182/ (accessed September 30, 2026). The committee-cited gnomAD record for p.Thr1515Met is https://gnomad.broadinstitute.org/variant/11-17394267-G-A?dataset=gnomad_r4. Dataset-specific frequency values were obtained from the committee review.

### Supplementary Table S3. Patient-linked non-yield findings retained for transparency

| Patient / sample ID | Gene, transcript annotation, and genomic position | Zygosity (GT) | Study reporting category | Reason for exclusion from primary outcome |
|---|---|---|---|---|
| FJ-ESRD-007 | *INF2*<br>NM_001031714:c.277T>G;<br>p.Ser93Ala<br>chr14:105167979 T>G (hg19) | Heterozygous (0/1) | Exploratory non-diagnostic finding | Gene–phenotype concordance, but sparse variant-specific clinical reports and insufficient segregation/functional evidence. Further well-characterized cases may support reassessment; no current diagnostic assignment. |
| FJ-ESRD-074 | *ABCC8*<br>NM_000352.6:c.4544C>T;<br>p.Thr1515Met<br>chr11:17415814 G>A (hg19) | Heterozygous (0/1) | Exploratory non-diagnostic finding | Committee VUS: PM1 + PM2 + PP3; PP4 not applied. Rare *NBD2* missense variant (REVEL 0.886). LP could be reached if PP4 were justified under an applicable framework; this remains conditional. Reassessment is appropriate as phenotype criteria or variant evidence evolve; no current diagnostic assignment. |
| FJ-ESRD-022 | *COQ8B*<br>NM_001142555: exon 1, 5′ UTR.<br>Location manually reviewed in IGV.<br>chr19:41220956 A>T (hg19) | Homozygous (1/1) | Exploratory non-diagnostic finding | Homozygous genotype and gene-disease plausibility, but the variant is noncoding. RNA-based functional analysis was not performed; its functional consequences remain unverified. |
| FJ-ESRD-005 | C3<br>NM_000064:c.1835C>T;<br>p.Thr612Met<br>chr19:6709705 G>A (hg19) | Heterozygous (0/1) | Exploratory non-diagnostic finding | Complement pathway relevance but insufficient evidence for patient-level diagnosis. |
| FJ-ESRD-086 | *CFHR5*<br>NM_030787:c.1303C>T;<br>p.Arg435Ter<br>chr1:196971767 C>T (hg19) | Heterozygous (0/1) | Exploratory non-diagnostic finding | Complement pathway relevance but insufficient evidence for patient-level diagnosis. |
| FJ-ESRD-088 | *COL4A3*<br>NM_000091:c.4349G>A;<br>p.Arg1450Gln<br>chr2:228172522 G>A (hg19) | Heterozygous (0/1) | Exploratory non-diagnostic finding | Collagen IV-related disease relevance but insufficient evidence for patient-level diagnosis. |

The six exploratory findings occurred in six distinct participants and did not overlap with the ten diagnoses shown in Table S4. All six findings were non-diagnostic and were excluded from the primary outcome numerator. They were not ranked as likely pathogenic, and the available rationale for retention and diagnostic limitations are stated for each finding.

*COQ8B* is an established CoQ10-related nephropathy gene.[S22] However, gene-level biological plausibility does not establish the effect of this 5′-UTR variant. The NM_001142555 location was manually reviewed in the IGV, and RNA-based analysis was not performed. Source annotations additionally include splicing/splice-5 for NM_001142555 and intronics for NM_024876, without an experimentally established splice effect.

*ABCC8* was reported using NM_000352.6. Transcript accession versions for the other findings (Table S3) and transcript-level cDNA HGVS for *COQ8B* were not supplied or inferred. Genomic REF/ALT alleles followed the supplied hg19 dataset, and cDNA substitutions followed the indicated transcript. GT, genotype; UTR, untranslated region; VUS, variant of uncertain significance.

### Supplementary Table S4. Patient-level summary of diagnoses after integrated clinical–genetic adjudication

Provided as a separate spreadsheet. The ten cases comprise nine P/LP-supported diagnoses and one clinically adjudicated Dent diagnosis with a *CLCN5* VUS. Seven diagnoses were renal whereas three were systemic/metabolic. The primary clinical–genetic outcome was 10/108 (9.3%; 95% CI, 5.1–16.2); the P/LP-supported subset excluding Dent was 9/108 (8.3%; 95% CI, 4.4–15.1). The *COL4A4* diagnosis in FJ-ESRD-061 was retained in the primary analysis and only omitted from the post hoc target-coverage sensitivity subset. Table S8 also reports the analyses excluding Gitelman syndrome. FJ-ESRD-044 clinical details include the absence of a personal or family history of neonatal hypoglycemia, insulin secretion, HbA1c, and treatment. Unavailable assay details and undocumented SGLT2 inhibitor exposure were separately identified.

#### FJ-ESRD-059: *SLC12A3*-related Gitelman syndrome

FJ-ESRD-059 carried homozygous *SLC12A3* NM_000339.3:c.2573T>A (p.Leu858His), a pathogenic variant associated with Gitelman syndrome (ClinVar Variation ID 225470). The pre-WGS clinical diagnosis was diabetic kidney disease. Health-check records from the patient’s twenties consistently showed serum potassium levels ≤3.5 mEq/L, with occasional values <3.0 mEq/L. The patient recalled episodes of polyuria, weakness, and fatigue beginning in the late teens. Earlier serum magnesium measurements were unavailable. However, the serum magnesium level was approximately 1 mEq/L after the development of kidney failure. Metabolic alkalosis could not be evaluated at our institution in the setting of kidney failure, and urinary calcium excretion was not assessed. The genotype and renal phenotype supported the Gitelman diagnosis, which was included among the seven renal diagnoses and in both the integrated and P/LP-supported outcome numerators. However, its contribution to kidney failure remains unclear. Table S8 shows an additional analysis omitting this diagnosis from the outcome numerators.

ClinVar: https://www.ncbi.nlm.nih.gov/clinvar/variation/225470/ (accessed September 30, 2026).

### Supplementary Table S5. Participant characteristics of the non-PKD WGS cohort

| Characteristic | Value | Available, n | Missing, n |
|---|---|---|---|
| Participants, n | 108 | 108 | 0 |
| Male, n (%) | 83 (76.9) | 108 | 0 |
| Female, n (%) | 25 (23.1) | 108 | 0 |
| Age at dialysis initiation, years | 40.5 (33.8–45.0) | 108 | 0 |
| Age at dialysis initiation, range | 20–50 | 108 | 0 |
| Age on June 1, 2024, years | 52.0 (46.0–57.0) | 108 | 0 |

Values are presented as medians (interquartile ranges), unless indicated otherwise. The percentage for sex include all 108 participants.

The age summaries used the confirmed census-date ages of all 108 participants. Sex was confirmed by the study investigators and was concordant with sequencing-based inference.

Family history frequencies were not tabulated because cohort-wide ascertainment and documentation were not sufficiently reliable. Individually verified family histories were reported in the case descriptions. The exact duration of dialysis was not derived from the integer age differences.

### Supplementary Table S6. Diagnoses by pre-WGS clinical category

The clinical categories were based on the pre-WGS records. The integrated diagnoses and the P/LP-supported subset are reported separately; the P/LP-supported subset excludes the clinically diagnosed case of Dent.

| Pre-WGS category | Enrolled, n | Integrated diagnoses, n/N (%) | P/LP-supported, n/N (%) |
|---|---|---|---|
| Diabetic kidney disease | 33 | 4/33 (12.1) | 3/33 (9.1) |
| Nephrosclerosis | 17 | 1/17 (5.9) | 1/17 (5.9) |
| Chronic glomerulonephritis | 19 | 2/19 (10.5) | 2/19 (10.5) |
| Unknown cause | 31 | 2/31 (6.5) | 2/31 (6.5) |
| Vasculitis | 1 | 0/1 (0.0) | 0/1 (0.0) |
| Other | 7 | 1/7 (14.3) | 1/7 (14.3) |
| **Total** | **108** | **10/108 (9.3)** | **9/108 (8.3)** |

FJ-ESRD-048 contributes to CGN, not unknown cause. FJ-ESRD-061 (FSGS) also contributes to CGN. The diagnosis in the Other category is FJ-ESRD-029 (interstitial nephritis before WGS). FJ-ESRD-059 retained the pre-WGS diabetic kidney disease category and contributed to the Gitelman diagnosis for both outcomes. Percentages use each row’s enrolled denominator, and categories are not interpreted as having equivalent detection rates. The integrated and P/LP-supported totals are 10/108 and 9/108, respectively.

### Supplementary Table S7. Nuclear candidate genes and supplied hg19 coordinates

Provided as a separate spreadsheet. The list contained 465 unique nuclear gene symbols with HGNC identifiers and hg19 coordinates. The Scope_and_sources and Source_literature sheets document investigator-reported curation, four source publications, and literature updates until July 2026. The coordinate-source labels were refGene_2020 for 460 genes and NCBI_RS2024_09 for *C4B*, *EBP*, *FANCD2*, *RPS17*, and *TNXB*, as supplied. The September 26, 2026, reporting snapshot is distinct from the original selection and analysis dates. Mitochondrial genes were not included in nuclear counts.

### Supplementary Table S8. Diagnostic proportions and post hoc target-coverage sensitivity analysis

Values are expressed as n/N, percentage, and the Wilson 95% confidence interval. The primary cohort comprised all 108 participants. The sensitivity analysis excludes FJ-ESRD-002, FJ-ESRD-003 and FJ-ESRD-061, each with <50% of the target bases covered at ≥10×, resulting in 105 participants.

| Diagnostic assessment | All participants<br>N = 108 | Post hoc target-coverage exclusion<br>N = 105 |
|---|---|---|
| Integrated clinical–genetic diagnoses | 10/108; 9.3%<br>(95% CI, 5.1–16.2) | 9/105; 8.6%<br>(95% CI, 4.6–15.5) |
| P/LP-supported diagnoses | 9/108; 8.3%<br>(95% CI, 4.4–15.1) | 8/105; 7.6%<br>(95% CI, 3.9–14.3) |
| Integrated diagnoses, excluding Gitelman | 9/108; 8.3%<br>(95% CI, 4.4–15.1) | 8/105; 7.6%<br>(95% CI, 3.9–14.3) |
| P/LP-supported diagnoses, excluding Gitelman | 8/108; 7.4%<br>(95% CI, 3.8–13.9) | 7/105; 6.7%<br>(95% CI, 3.3–13.1) |

The P/LP-supported subset excluded the phenotype-based clinical Dent diagnosis associated with a *CLCN5* VUS. The same target coverage rule excluded FJ-ESRD-002, FJ-ESRD-003, and FJ-ESRD-061 irrespective of the diagnosis. Because FJ-ESRD-061 contributes to the *COL4A4* diagnosis, the integrated numerator decreases from 10 to 9 and the P/LP-supported numerator from 9 to 8; the FJ-ESRD-061 diagnosis remains in the primary analysis. The last two rows additionally omit the Gitelman diagnosis in FJ-ESRD-059 from the numerators, while retaining the participant in the denominators. The contribution of Gitelman syndrome to kidney failure remains unclear. These post-hoc analyses did not validate the sensitivity of WGS. The calculations and reported low-coverage subset are provided in the spreadsheet of Supplementary Table S8. Supplementary Table S10 shows the complete 108-participant Per_sample QC table.

### Supplementary Table S9. Selected CNV/SV findings and patient-level interpretation

Provided as a separate spreadsheet. The table records the computational support for the counted NPHP1 diagnosis and selected non-diagnostic findings involving *ALPL*, *AGT*, *BBS5*, *RNF213*, *APOL1* vicinity, and intronic *PKHD1* gain in FJ-ESRD-067. These findings did not establish any additional diagnoses of kidney failure. The *PKHD1* signal at hg19 chr6:51,827,912–51,867,909 spanned four CNVkit bins (log2 +0.26), spared coding exons, and lacked Manta support. The *BBS5*/*AGT* sequence review is limited to available coding-variant calls, and non-explanatory findings are not automatically classified as benign.

The CNV/SV findings were interpreted for 98 of the 108 participants. This denominator is distinct from that of the 105-participant target-coverage sensitivity cohort. Participant-level QC and confirmed study-to-technical ID links are shown in Supplementary Table S10.

### Supplementary Table S10. Per-participant sequencing QC

Provided as a separate spreadsheet. The Per_sample sheet contains one row per confirmed participant, with study and technical identifiers, nuclear and mitochondrial depth metrics, target-coverage fractions, investigator-confirmed sex, and analysis-specific inclusion flags. The original QC measurements and classifications were preserved. All summaries in this table use the same 108 participants.

| Metric | n or median | Range / scope |
|---|---|---|
| Participants | 108 | Confirmed study cohort |
| CNV/SV evaluable | 98 | 92 standard + 6 batch reference |
| Target-coverage sensitivity cohort | 105 | FJ-ESRD-002, FJ-ESRD-003 and FJ-ESRD-061 excluded |
| Genome-wide mean depth, × | 13.7 | 3.8–61.1 |
| Target-region mean depth, × | 14.2 | 5.7–59.5 |
| Target bases covered at ≥10×, % | 86.9 | 16.9–99.9 |
| Autosomal target bases covered at ≥10×, % | 89.0 | 17.5–99.9 |
| Target bases covered at ≥20×, % | 10.5 | 0.2–99.5 |
| mtDNA mean coverage (MitoHPC), × | 1,005.8 | 422.3–1,883.1 |

Depth and coverage summaries are expressed as medians of sample-level values; they are not validation estimates of variant-detection sensitivity. CNV/SV evaluability and the coverage-based sensitivity analyses were performed separately. Sex was confirmed by the study investigators and was concordant with the sex chromosome read-depth inference.

### Supplementary References


S1. Granhøj J, Aagaard MM, Pedersen KV, et al. Diagnostic yield of whole-genome sequencing in patients with kidney failure of undetermined etiology at age 50 years or younger. Kidney Int Rep. 2025;10(11):3984–3997. doi:10.1016/j.ekir.2025.09.001.

S2. Saei H, Morinière V, Heidet L, et al. VNtyper enables accurate alignment-free genotyping of MUC1 coding VNTR using short-read sequencing data in autosomal dominant tubulointerstitial kidney disease. iScience. 2023;26(7):107171. doi:10.1016/j.isci.2023.107171.

S3. Pedersen BS, Quinlan AR. Mosdepth: quick coverage calculation for genomes and exomes. Bioinformatics. 2018;34(5):867–868. doi:10.1093/bioinformatics/btx699.

S4. Talevich E, Shain AH, Botton T, Bastian BC. CNVkit: genome-wide copy number detection and visualization from targeted DNA sequencing. PLoS Comput Biol. 2016;12(4):e1004873. doi:10.1371/journal.pcbi.1004873.

S5. Chen X, Schulz-Trieglaff O, Shaw R, et al. Manta: rapid detection of structural variants and indels for germline and cancer sequencing applications. Bioinformatics. 2016;32(8):1220–1222. doi:10.1093/bioinformatics/btv710.

S6. Battle SL, Puiu D, Verlouw J, et al. A bioinformatics pipeline for estimating mitochondrial DNA copy number and heteroplasmy levels from whole genome sequencing data. NAR Genom Bioinform. 2022;4(2):lqac034. doi:10.1093/nargab/lqac034.

S7. Weissensteiner H, Forer L, Fendt L, et al. Contamination detection in sequencing studies using the mitochondrial phylogeny. Genome Res. 2021;31(2):309–316. doi:10.1101/gr.256545.119.

S8. Groopman EE, Marasa M, Cameron-Christie S, et al. Diagnostic utility of exome sequencing for kidney disease. N Engl J Med. 2019;380:142–151. doi:10.1056/NEJMoa1806891.

S9. Chebib FT, Wang X, Udani SM, et al. Genetic testing in the management of adult CKD. J Am Soc Nephrol. 2026;37(4):777–789. doi:10.1681/ASN.0000000913.

S10. Mallawaarachchi AC, Fowles L, Wardrop L, et al. Genomic testing in patients with kidney failure of an unknown cause: a national Australian study. Clin J Am Soc Nephrol. 2024;19(7):887–897. doi:10.2215/CJN.0000000000000464.

S11. Doreille A, Lombardi Y, Dancer M, et al. Exome-first strategy in adult patients with CKD: a cohort study. Kidney Int Rep. 2023;8(3):596–605. doi:10.1016/j.ekir.2022.12.007.

S12. Richards S, Aziz N, Bale S, et al. Standards and guidelines for the interpretation of sequence variants: a joint consensus recommendation of the American College of Medical Genetics and Genomics and the Association for Molecular Pathology. Genet Med. 2015;17:405–424. doi:10.1038/gim.2015.30.

S13. Savige J, Storey H, Watson E, et al. Consensus statement on standards and guidelines for the molecular diagnostics of Alport syndrome: refining the ACMG criteria. Eur J Hum Genet. 2021;29:1186–1197. doi:10.1038/s41431-021-00858-1.

S14. Pejaver V, Byrne AB, Feng BJ, et al. Calibration of computational tools for missense variant pathogenicity classification and ClinGen recommendations for PP3/BP4 criteria. Am J Hum Genet. 2022;109(12):2163–2177. doi:10.1016/j.ajhg.2022.10.013.

S15. Bökenkamp A, Ariceta G, Böckenhauer D, et al. Dent disease: clinical practice recommendations. Nephrol Dial Transplant. 2025;40:852–864.

S16. Boodhansingh KE, Kandasamy B, Mitteer L, et al. Novel dominant KATP channel mutations in infants with congenital hyperinsulinism: validation by in vitro expression studies and in vivo carrier phenotyping. Am J Med Genet A. 2019;179:2214–2227. doi:10.1002/ajmg.a.61335.

S17. MacMullen CM, Zhou Q, Snider KE, et al. Diazoxide-unresponsive congenital hyperinsulinism in children with dominant mutations of the β-cell sulfonylurea receptor SUR1. Diabetes. 2011;60:1797–1804. doi:10.2337/db10-1631.

S18. Scala R, Li Y, Gao J, et al. Paradoxical maturity-onset diabetes of the young arising from loss-of-function mutations in ATP-sensitive potassium channels. Diabetes. 2026;75:180–192. doi:10.2337/db25-0110.

S19. Yoshiji S, Hasebe M, Tanaka D, et al. Genetic and clinical characteristics of monogenic diabetes in Japan: a nationwide study by the Japan Diabetes Society. J Clin Endocrinol Metab. 2026;111:757–769. doi:10.1210/clinem/dgaf478. Published online August 25, 2025.

S20. Clemente M, Cobo P, Antolín M, et al. Genetics and natural history of non-pancreatectomized patients with congenital hyperinsulinism due to variants in ABCC8. J Clin Endocrinol Metab. 2023;108:e1316–e1328. doi:10.1210/clinem/dgad280.

S21. Kapoor RR, Flanagan SE, James CT, et al. Hyperinsulinaemic hypoglycaemia and diabetes mellitus due to dominant ABCC8/KCNJ11 mutations. Diabetologia. 2011;54:2575–2583. doi:10.1007/s00125-011-2207-4.

S22. Ashraf S, Gee HY, Woerner S, et al. ADCK4 mutations promote steroid-resistant nephrotic syndrome through CoQ10 biosynthesis disruption. J Clin Invest. 2013;123:5179–5189.
